# Machine learning-based proteogenomic data modeling identifies circulating plasma biomarkers for early detection of lung cancer

**DOI:** 10.1101/2024.07.30.24311241

**Authors:** Marcela A. Johnson, Liping Hou, Bevan Emma Huang, Assieh Saadatpour, Abolfazl Doostparast Torshizi

## Abstract

Identifying genetic variants associated with lung cancer (LC) risk and their impact on plasma protein levels is crucial for understanding LC predisposition. The discovery of risk biomarkers can enhance early LC screening protocols and improve prognostic interventions. In this study, we performed a genome-wide association analysis using the UK Biobank and FinnGen. We identified genetic variants associated with LC and protein levels leveraging the UK Biobank Pharma Proteomics Project. The dysregulated proteins were then analyzed in pre-symptomatic LC cases compared to healthy controls followed by training machine learning models to predict future LC diagnosis. We achieved median AUCs ranging from 0.79 to 0.88 (0-4 years before diagnosis/YBD), 0.73 to 0.83 (5-9YBD), and 0.78 to 0.84 (0-9YBD) based on 5-fold cross-validation. Conducting survival analysis using the 5-9YBD cohort, we identified eight proteins, including CALCB, PLAUR/uPAR, and CD74 whose higher levels were associated with worse overall survival. We also identified potential plasma biomarkers, including previously reported candidates such as CEACAM5, CXCL17, GDF15, and WFDC2, which have shown associations with future LC diagnosis. These proteins are enriched in various pathways, including cytokine signaling, interleukin regulation, neutrophil degranulation, and lung fibrosis. In conclusion, this study generates novel insights into our understanding of the genome-proteome dynamics in LC. Furthermore, our findings present a promising panel of non-invasive plasma biomarkers that hold potential to support early LC screening initiatives and enhance future diagnostic interventions.

## INTRODUCTION

Lung cancer (LC) is the leading cause of cancer related deaths worldwide. Recent projections in the US for 2024 estimate 200,000 newly diagnosed cases and over 100,000 mortalities (*1*). Given the advancements in early detection, reductions in smoking, and novel therapeutic interventions, the mortality rate of LC has declined. However, despite significant advancements, the 5-year overall survival rate for LC remains between 25-63%, depending on the stage at diagnosis. Almost three-quarters of cases are still diagnosed in late stages, highlighting the unmet need for enhanced early detection interventions to improve the disease outcomes (*2*). The criteria established by the US Preventive Service Task Force recommends low dose computed tomography screening for high-risk population (*3*). The high-risk population includes those who are current smokers or have quit in the past 15 years, are between 50-80 years, and have a smoking history of 20 pack-years (*3*). However, this high-risk classification only encompasses 4% of the total US population (*2*). Although smoking remains the biggest risk factor, it has been estimated that about 15-20% of worldwide cases occur among non-smokers^2^ and that about 18% of LC cases are heritable (*4–6*). Therefore, there is a crucial need to discover minimally invasive biomarkers to incorporate genetic predisposition and exposure to environmental factors and expand the population of high-risk individuals eligible for early screening of LC (*7*).

Genome-wide association studies (GWAS) have identified over 40 loci associated with LC susceptibility (*8*). While many such loci have been functionally evaluated beyond association (*8–12*), the etiology of LC risk remains underexplored. Protein dynamics are intricately linked to the genetic architecture of humans and serve as essential components of cellular machinery. Genetic aberrations have the potential to significantly impact the normal functionality of proteins, thereby increasing the risk of developing various diseases (*13, 14*). Circulating plasma proteins serve as reliable indicators of the physiological state of various tissues and organs and offer valuable insights into the alterations occurring in affected tissues as well as the systemic changes associated with a particular disease. Moreover, due to the non-invasive nature of blood sampling, plasma proteins are easily accessible for evaluating disease-related complications. Integrating GWAS and proteome enables a comprehensive understanding of the interplay between the human genetic architecture and proteome dynamics, shedding light on disease predisposition and uncovering potential biomarkers or pathways associated with diseases (*14–19*).

Population-based biobanks have played a crucial role in uncovering genetic variants associated with numerous phenotypes, allowing the integration of health and lifestyle factors into GWAS. The UK Biobank (UKB), which has characterized the genetic profiles of about 500,000 individuals and includes longitudinal electronic health records, offers a valuable resource in this regard (*20*). Large-scale proteomic studies have recently been revolutionized by the development of high-throughput multiplex affinity proteomics technologies (*21*). Previous research has demonstrated the significance of exploring the associations between genetic variants and plasma protein levels (pQTLs) in various diseases, including LC (*12, 22–26*). However, a major limitation of these studies has been the absence of longitudinal data and comprehensive phenotypic, health, and lifestyle information for each participant. The UK Biobank Pharma Proteomics Project (UKB-PPP), coupling rich phenotypic and genomic data, offers an unprecedented opportunity to overcome this limitation (*18*). The UKB-PPP includes 2,923 proteins measured across 54,219 UKB participants using the antibody-based proximity extension assay (*18*).

Several studies have examined the potential applications of plasma proteins as biomarkers for the future diagnosis of LC. Many of these studies have been constrained either because of a limited set of proteins or to a time window of up to five years from sample collection to diagnosis (*18, 27, 28*).

However, considering that LC is frequently asymptomatic in its early stages or exhibits non-specific symptoms, it is crucial to investigate whether plasma proteins can detect the risk of LC even earlier than five years prior to diagnosis (*29*). Such research holds significance in enhancing disease prognosis and advancing early detection strategies for improved patient outcomes.

Thus, in this study, we leveraged data from the UKB and other large-scale population genetic studies to characterize genetic variants associated with LC risk and the levels of plasma proteins. By screening plasma proteins from the UKB-PPP, we identified potential biomarkers indicative of a future diagnosis of LC within a 9-year timeframe following sample collection. These findings contribute to our understanding of the relationship between genetic factors, proteomic profiles, and the development of LC, and to identify novel potential biomarkers to support the screening criteria improving early detection and intervention strategies for this disease.

## RESULTS

To identify potential biomarkers associated with lung cancer, a two-stage strategy was implemented. In the first stage, three separate analyses were performed. LC cases and controls from the UKB were utilized to conduct a GWAS to investigate the genetic susceptibility of LC (UKB GWAS). A meta-analysis was conducted combining the summary statistics from the UKB GWAS with the summary statistics published by FinnGen (*30*). Lastly, to identify the loci associated with both LC and changes in plasma protein levels, we integrated the findings from the GWAS analyses, and the genome-wide significant variants (Bonferroni correction threshold of *P* − *value* < 5*e*^-8^) reported in the meta-analysis conducted by McKay *et al.* (*31*). This integration allowed us to optimize the characterization of UKB-PPP protein quantitative trait loci (pQTLs) in LC context (Fig. 1).

**Fig. 1.**
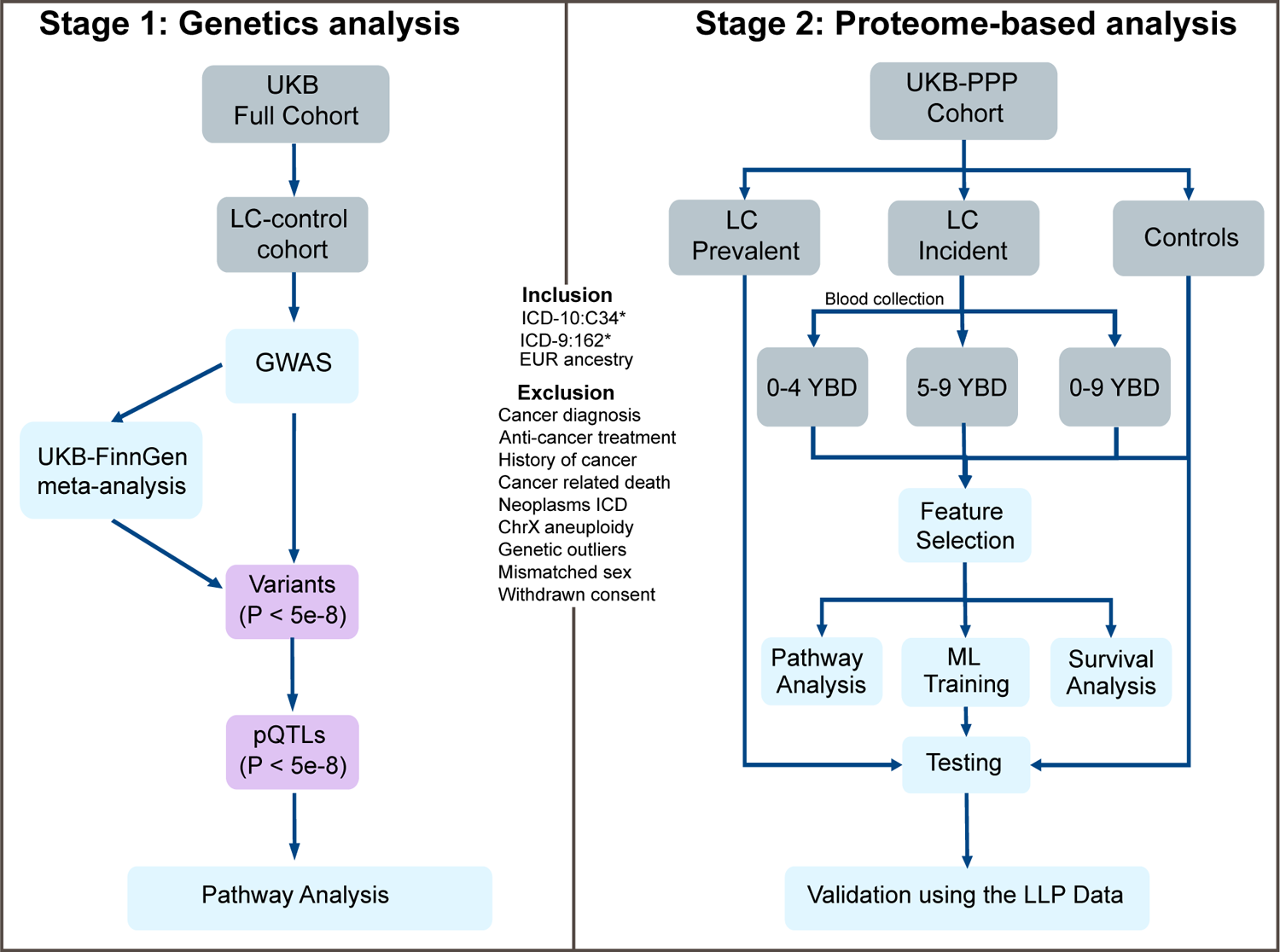
Study design. Workflow depicting the comprehensive two-stage study design integrating data from the UKB and UKB-PPP. The study design encompassed standardized inclusion and exclusion criteria, consistently applied in both stages.

In the second stage, we utilized the data from the UKB-PPP to categorize participants into three different groups: incident cases (those who developed lung cancer after blood collection), prevalent cases (those already diagnosed with lung cancer at the time of sample collection), and controls (individuals without an LC diagnosis during the follow up period and no personal history of any cancer type). To analyze the relationship between protein levels and different LC development time frames, we divided the incident cases into three subgroups based on the time from sample collection to diagnosis: 0-4 years before diagnosis (YBD), 5-9 YBD, and 0-9 YBD. We compared the protein levels across the different cohorts to identify proteins that differentiate LC cases from controls. To gain deeper insights into the underlying biology and potential mechanisms associated with the selected proteins, we conducted pathway enrichment analysis followed by investigating the impact of protein levels on the overall survival of patients. The selected proteins were used as predictive features to train six machine learning models and create a concise panel of potential biomarkers that can predict the risk of a future LC diagnosis.

Finally, to validate the robustness of the selected proteins, we performed an external validation using a recently published cohort from the Liverpool Lung Project (LLP) (*32*) (Fig. 1).

### Genetics Analysis

#### GWAS using UKB-derived cohort

To identify variants associated with overall lung cancer, we conducted a GWAS utilizing UKB. We created a cohort of 4,892 individuals of European (EUR) ancestry diagnosed with LC during the follow up time, referred to as LC cases, and 260,275 individuals with no history of cancer, referred to as controls. The median age of the cohort at sampling was 63 for cases and 57 for controls. Among the participants, approximately 48% of LC cases and 53% of controls were female, and 83% of LC cases and 43% of controls were current or former smokers (Table 1).

**Table 1.**
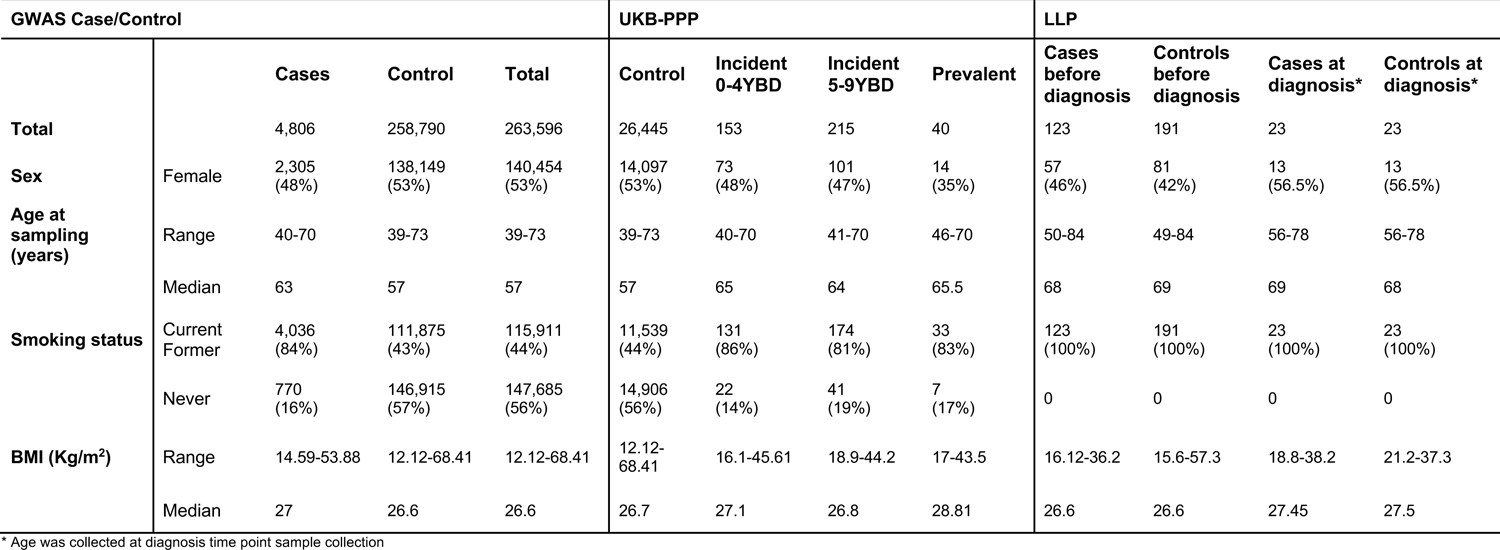
Demographics of the cohorts used in this study.

We performed a meta-analysis using the UKB GWAS together with the summary statistics published by FinnGen (*30*), with a cohort of 12,303 LC cases and 572,983 controls (see Materials and Methods). We identified 5 significant loci (*P* − *value* < 5*e*^-8^) in the UKB GWAS and 10 additional loci harboring significant variants in the UKB-FinnGen meta-analysis (Fig. 2 and Table S1 to S3). Of these 15 loci, ten had been previously reported in a GWAS (*9*) performed with a different dataset, while the remaining five had not previously reached genome-wide significance (*11, 31*) (Fig. 2). Among these novel loci, all sentinel variants (the most significant SNPs) were found to be in intronic regions. Based on RegulomeDB (RDB) scores (*33*), four of these sentinel variants were found to have putative functional roles. For example, the variant rs2524296 in the 11q12.2 locus near the *FADS1* gene, shows a RDB score of 1f (Table S2). This variant is an expression quantitative trait locus (eQTL) for four different genes, including *FADS1* in the upper lobe of the left lung, and influences the binding of 12 transcription factors, indicating a potential regulatory role in gene expression (*31*). It is important to note that a different variant (rs174559), within the same locus, has recently been reported to be associated with lung adenocarcinoma (LUAD) and colocalized with a lung eQTL in individuals of East Asian ancestry, but has not been reported to be associated with LC in individuals of EUR ancestry (*11*). Lower expression of *FADS1* has also been associated with poorer overall survival and disease-free survival in non-small cell lung cancer (NSCLC) patients (*34*). The loci 10q24 (rs9419958) and 20q13 (rs6011779) loci were also found to be associated with LC in the latest meta-analysis published by McKay *et al.* (*31*) (Fig. 2B and Table S2). The sentinel variants in 10q24 are in proximity to the *OBFC1/STN1* gene (Fig. 2B and Table S2).

**Fig. 2.**
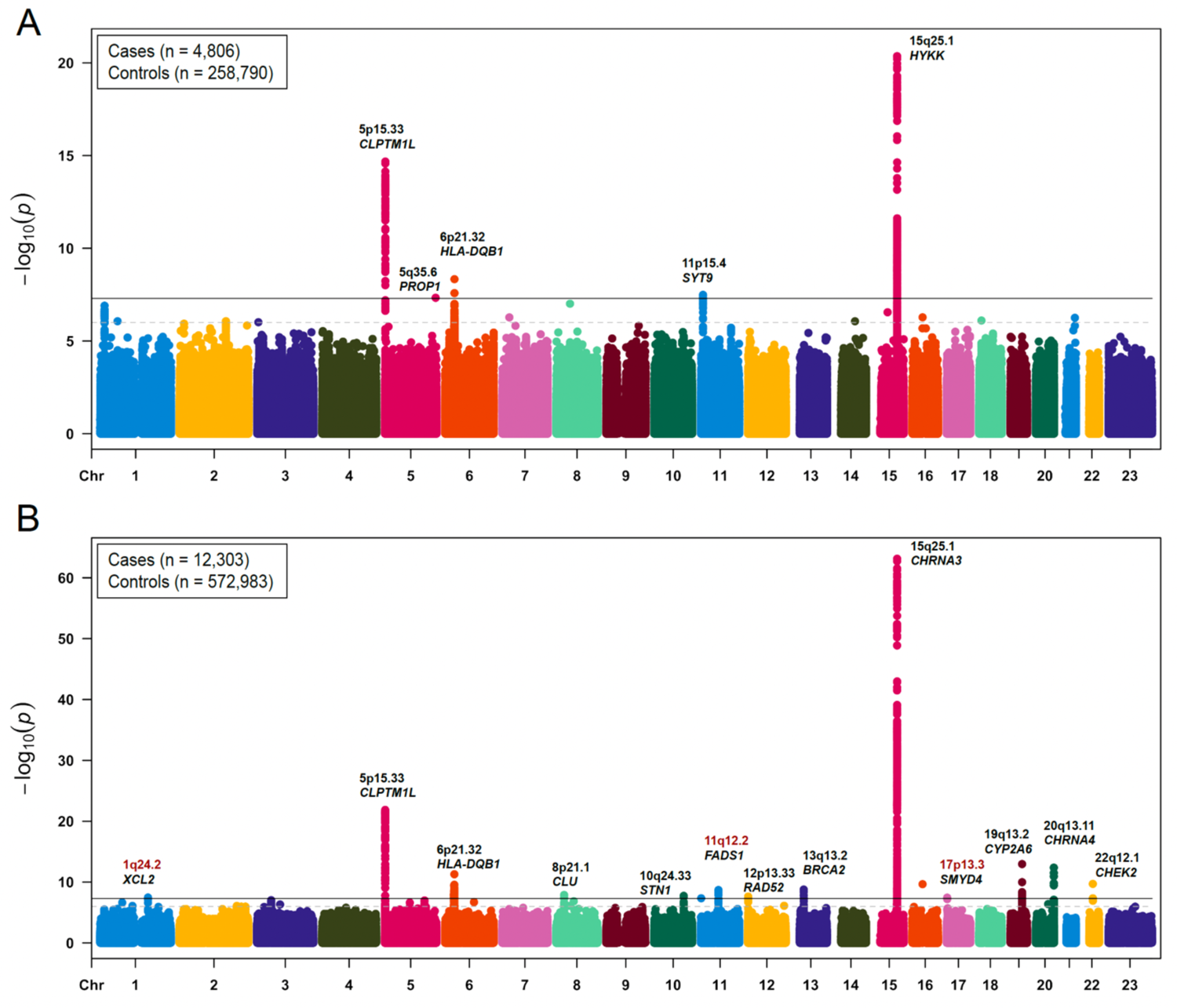
Manhattan plots illustrating the results from both GWAS performed. A) UKB LC cohort B) Meta-analysis conducted using the summary statistics from the UKB GWAS and FinnGen. In both, variants that achieved genome-wide significance (p-value < 5e-8) are labeled. Previously associated variants with lung cancer are annotated in black and other variants are depicted in red.

#### UKB-PPP pQTLs associated with LC

To investigate potential associations between the levels of proteins detected in plasma and the significant variants in the UKB GWAS, UKB-FinnGen and the McKay *et al.* (*31*) GWAS, we utilized the recently characterized dataset of plasma pQTLs (*18*) to identify pQTLs associated with both traits.

From our analysis, we identified 166 variants from the UKB GWAS, 511 variants from the UKB-FinnGen meta-analysis, and 1,464 variants from the McKay *et al.* (*31*) meta-analysis that are also significant pQTLs (*P* − *value* < 5*e*^-8^) in plasma (Fig. 3A). These significant pQTLs were associated with a total of 638 unique proteins. Among these proteins, four were exclusively associated with variants from the UKB GWAS, 32 with the UKB-FinnGen meta-analysis, 163 with the variants reported in the McKay *et al.* (*31*) meta-analysis, and 138 were found in all the analyses (Fig. 3B and Table S4).

**Fig. 3.**
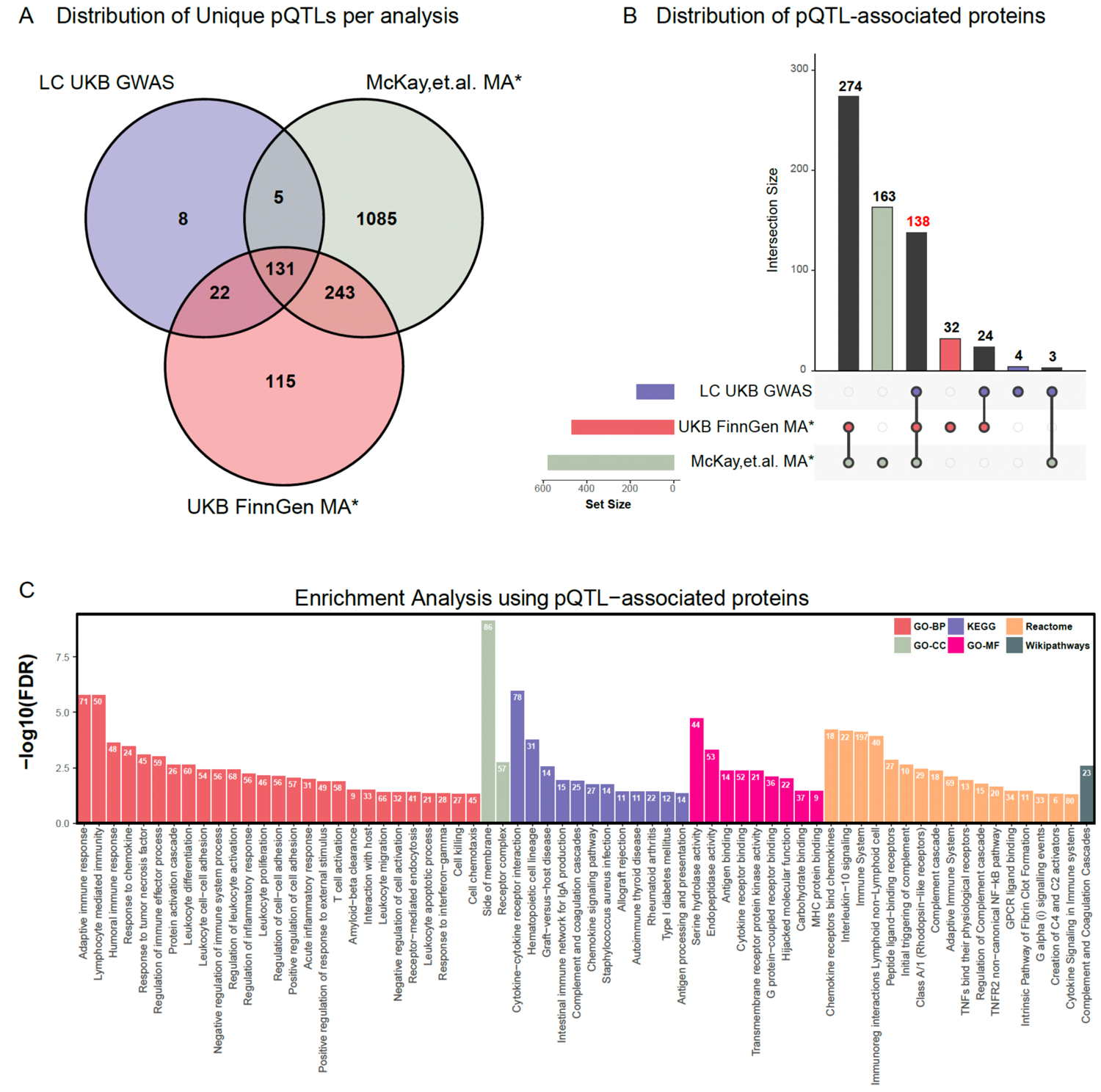
Downstream analysis of pQTL-associated proteins. A) Distribution and intersections of the unique pQTLs among different studies are depicted using a Venn Diagram. B) The UpSet plot showcases the overlap of pQTL-associated proteins across the various studies. Each colored bar represents the number of proteins found in the corresponding study. The numbers on top of the vertical bars indicate the protein count in each intersection, with the number in red highlighting the proteins found in all. *MA stands for meta-analysis. C) Enrichment analysis for Gene Ontology (GO), KEGG, Reactome, and Wikipathways of the pQTL-associated proteins using WebGestalt (https://www.webgestalt.org/), with the UKB-PPP proteins serving as the background gene set. All displayed pathways achieved statistical significance with a false discovery rate (FDR) threshold of less than 0.05. The number above each bar represents the observed protein frequency associated with each pathway.

To examine the relationship between the identified pQTLs and the genes encoding the target proteins, we defined a *cis* association as the pQTL localized within 1Mb window flanking the gene encoding the protein of interest. Otherwise, the association is called *trans* (Materials and Methods). Overall, the *cis*-pQTLs were associated with the expression levels of proteins EPHX2, BRSK2, SCN4B, TREH, TMEM25, and CTSH. It is worth highlighting that the *cis*-pQTLs (rs11852372, rs12914385, and rs55781567) linked to higher protein levels in plasma of CTSH coincided with the sentinel variants that demonstrated positive associations LC in the UKB GWAS and both meta-analyses (Table S4). Several biological pathways found to be related to the significant pQTL-associated proteins including the immune system pathway (*FDR* = 8*e*^-5^), cytokine-cytokine receptor interaction (*FDR* = 1*e*^-5^), GPCR ligand binding (*FDR* = 0.0012), adaptive immune response (*FDR* = 6*e*^-8^), and endopeptidase activity (*FDR* = 3*e*^-5^). The panel used for the UKB-PPP dataset is enriched for proteins associated with immune-related diseases, therefore a high enrichment of immune-related pathways was expected (Fig. 3C and Table S6).

#### Proteomics Analysis UKB-PPP LC cohort

To identify proteins associated with a future diagnosis of LC, we utilized different sets of participants from the UKB-PPP. These patient sets included 153 participants who were diagnosed with LC less than 5 years post blood collection (0-4 YBD), 215 patients with a diagnosis of LC more than 5 and up to 9 years post blood donation (5-9 YBD), 40 prevalent cases, and 26,445 controls without any history of cancer (Table 1 and Fig. 4A).

**Fig. 4.**
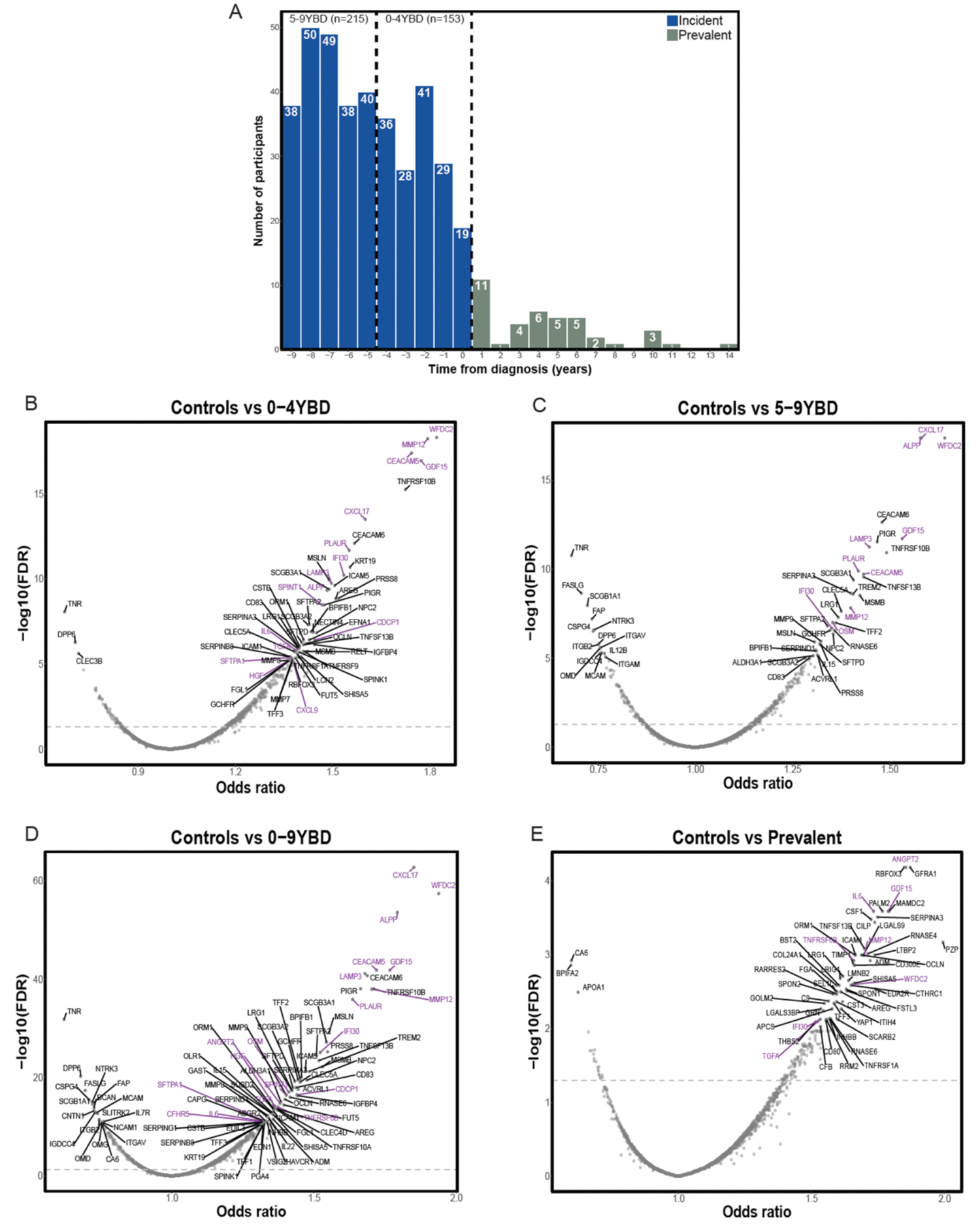
UKB-PPP data and DE analysis. A) Bar plot showing the distribution of the LC cases in the UKB-PPP. The cases are categorized into two groups: incident cases and prevalent cases. Incident cases represent individuals who were diagnosed with LC after blood sample collection, while prevalent cases include individuals who already had a confirmed LC diagnosis at the time of blood collection. B-E) DE analysis of the different categories against controls. The significance threshold was set at FDR < 0.05 (dashed line). The proteins labeled in purple represent those that have previously been reported as potential biomarkers by the LC3 study.

#### Differentially expressed plasma proteins in participants with a future LC diagnosis

After filtering the proteins measured by the UKB-PPP, we used 2,937 analytes capturing 2,919 unique proteins for conducting the differential expression analyses (Materials and Methods). When using the 0-4YBD cohort to obtain proteins that differentiate incident cases and controls, we obtained 410 differentially expressed (DE) analytes, out of which 347 were up-regulated (with odds ratio (OR) > 1) and 63 were down-regulated (OR < 1) (Fig. 4B). In the 5-9YBD cohort, we identified 340 DE analytes, with 243 being up-regulated and 97 being down-regulated (Fig. 4C). Furthermore, in the 0-9YBD cohort, we identified 807 DE analytes, with 594 up-regulated and 213 down-regulated. Lastly, comparing prevalent cases with controls yielded 193 DE analytes where 172 were up-regulated and 21 were down-regulated (Fig. 4E).

#### Feature selection, predictive modeling, and optimal features

Through the implementation of a bootstrapping strategy aimed at acquiring a robust set of features for predictive modeling, we assessed the stability and consistency of the analytes in their capacity to effectively differentiate between cases and controls (Materials and Methods). Based on this analysis, we identified 869 DE analytes (*FDR* < 0.05 in at least 100 out of 1000 bootstrapping iterations) within the 0-4YBD cohort, 782 DE analytes within the 5-9 YBD cohort, and 491 DE analytes within the 0-9YBD cohort (Table S5).

To evaluate the predictive power of the selected analytes and ML models in accurately classifying LC cases in each of the cohorts, we used a 5-fold cross validation (CV) approach. During CV, we trained six ML models: Logistic Regression (LogReg), Elastic Net Regression (ElasticNet), Extreme Gradient Boosting (XGBoost), Support Vector Machine (SVM), Random Forest (RF), and a feed forward three-layer Neural Network (NN) with 100 neurons in the hidden layer. The performance of the models was measured using the Area Under the Receiver Operating Characteristic Curve (AUC) and F1 scores (Materials and Methods).

Using the 869 features selected using the 0-4YBD cohort, the median AUC scores for the models ranged between 0.79-0.88 (Fig. 5A). Similarly, with the 782 selected features using the 5-9YBD cohort, the median AUC ranged from 0.73 to 0.83 (Fig. 5B). Furthermore, using the 491 selected features using the 0-9YBD cohort, the median AUC values ranged between 0.78 and 0.84 (Fig. 5C). To evaluate the effectiveness of the models and the selected features in classifying diagnosed LC cases, as a test set, we utilized the prevalent cases along with the 40 participants from the control cohort that were not included in the training (Materials and Methods). Among the models trained on the different cohorts, RF demonstrated the best performance with AUC values of 0.81 for 0-4YBD (F1 score of 0.81), 0.85 for 5-9YBD (F1 score of 0.85), and 0.84 for 0-9YBD (F1 score of 0.84) (Fig. 5D-F). To address potential correlations among the selected features and identify proteins as potential biomarkers, we performed a recursive feature elimination with cross-validation (RFECV) applied to RF (Materials and Methods). The number of optimal features obtained were 209 for 0-4YBD, 47 for 5-9YBD, and 71 for 0-9YBD. When comparing the different optimal features, we noticed that 22 proteins were selected in all the cohorts. Out of these commonly selected features, eight proteins, CEACAM6, FASLG, TNR, EDA2R, NPC2, IGDCC4, LRG1, and ASGR2, have not yet been reported as potential biomarkers for early detection of LC (Table S8).

**Fig. 5.**
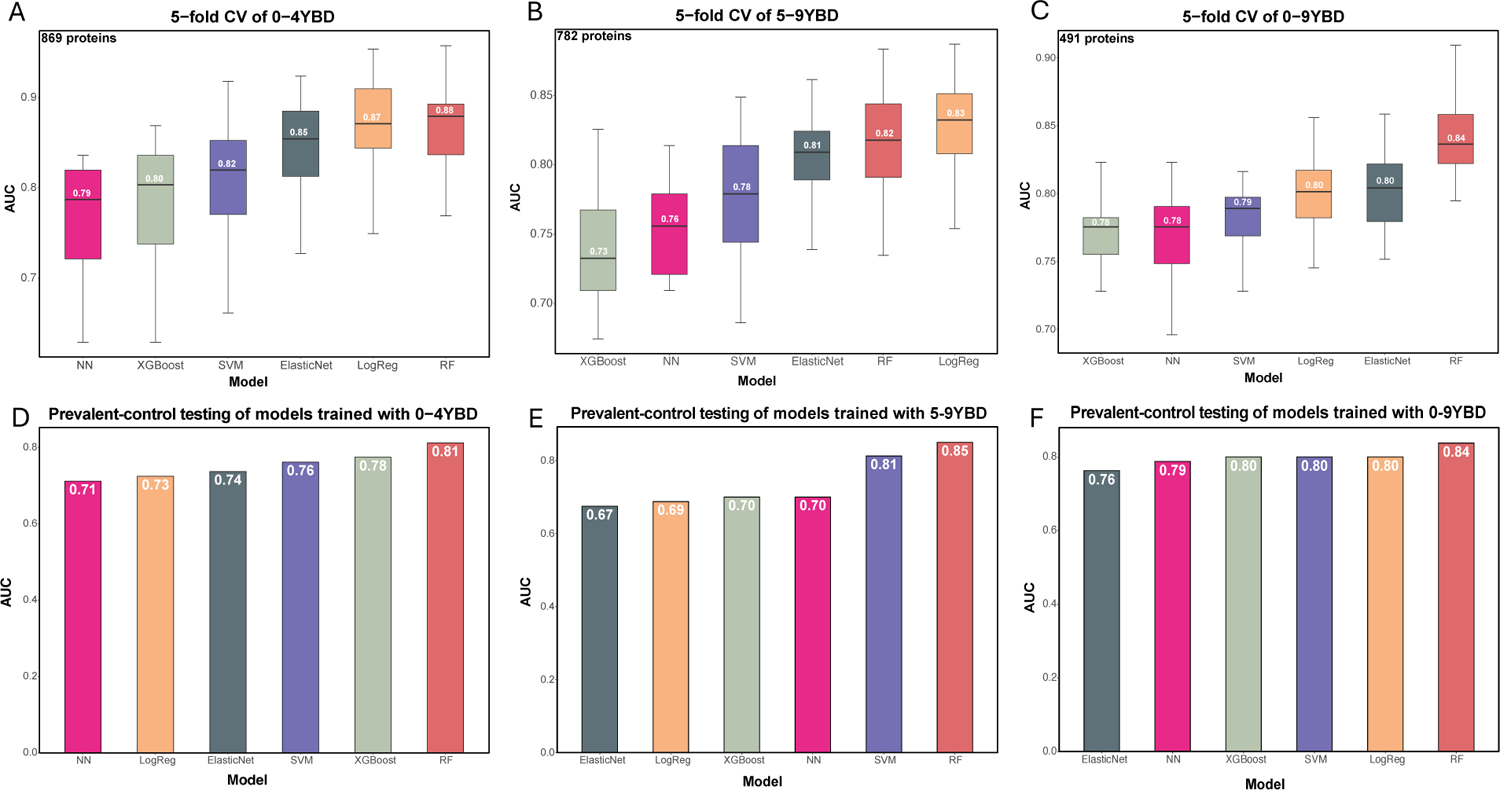
Predictive modeling of LC using circulating plasma proteins. A-C) A boxplot depicting the AUC values from the 5-fold CV repeated 5 times using the six different machine learning models: Logistic Regression, Random Forest, Elastic Net, Support Vector Machine, XGBoost, and Neural Network. The analysis employed 869, 782, and 491 proteins in conjunction with the corresponding incident cohorts. D-F) Bar plots representing the AUC values of the models when tested with the prevalent and control cohorts. The AUC values serve as a measure of predictive performance for the models.

#### Validation of selected features using cohorts from the Lung Liverpool Project (LLP)

In the study published by Davies *et al*. (*32*), the authors used a case-control cohort from the LLP, a longitudinal observational study that includes samples taken before and at the diagnosis. To evaluate the generalizability and robustness of the features selected from the UKB-PPP cohorts, we used this cohort from the LLP, including samples taken up to 9 years before diagnosis, samples taken at diagnosis time point, and healthy controls (Table 1).

The median age for the LLP cohorts was generally higher (68 and 69) compared to the median age in the UKB-PPP cohorts (65 for 0-4YBD and prevalent cases, 64 for 5-9YBD incident cases and 57 for all controls). The percentage of females was higher in the LLP diagnosis cohort compared to the UKB-PPP prevalent cases (56% vs. 35%), and all the LLP participants were smokers (Table 1). We assessed the performance of the six ML models using three different LLP cohorts based on the time to diagnosis, with the cohort from the diagnosis time point serving as the testing set (Materials and Methods). When testing on individuals who were diagnosed with LC less than 5 years after blood collection (1-5Y) or up to 9 years after blood collection (1-9Y), along with controls, the SVM model exhibited the highest performance with AUCs of 0.78 and 0.80, respectively. In contrast, when testing on individuals diagnosed with LC between 5 and less than 9 years (5-9Y) after blood collection, the ElasticNet model achieved the highest performance with an AUC of 0.67 (Fig. S2). It is noteworthy that minimal differences in protein expression between controls and cases were observed in the 5-9Y category, which likely impacted the results observed in this validation. It is important to mention that due to the lack of cancer staging information in the UKB-PPP, and to mitigate the inclusion of individuals with undiagnosed diseases, only the 5-9YBD cohort was included for further analysis.

#### Downstream analysis of the 5-9YBD selected proteins

To assess the potential biological functions and pathways associated with the development of LC, we conducted an enrichment analysis using the proteins selected from the 5-9YBD cohort (Materials and Methods). The enrichment analysis revealed several significant biological pathways including leukocyte migration of biological process, extracellular matrix of cellular components, receptor ligand activity in molecular function, the immune system pathway, and the cytokine-cytokine receptor interaction pathway (Fig. S1). It is worth noting that previous studies have demonstrated the significant role of cytokines in LC immunity, as they can either promote or inhibit tumor growth^17^, which further supports the relevance of the observed cytokine-cytokine receptor interaction pathway in the context of LC development.

Using the selected proteins, we investigated the influence of protein expression on patient survival within the 5-9YBD cohort, utilizing a Cox proportional-hazard model (Materials and Methods). Out of the 215 cases, 72% experienced death events during the follow-up period resulting in a 5-year survival rate of 13.5% and a median survival of 13.5 months. Among the top 25 proteins, the expression levels of seven proteins, EDA2R, CHRDL1, CD74, TNFRSF21, ACVRL1, COL6A3, and CALCB, were significantly associated with a lower survival rate (HR>1.3 and FDR<0.05) (Table S7). We performed a survival analysis using the Kaplan-Meier estimator to investigate the association of expression of the seven proteins and PLAUR/uPAR, which has been reported to be a prognostic biomarker of LC (*35*), with overall survival (OS) (Materials and Methods). The lower expression of all eight proteins was significantly associated (*P* − *value* < 0.05 log-rank test) with better survival (Fig. 6).

**Fig. 6.**
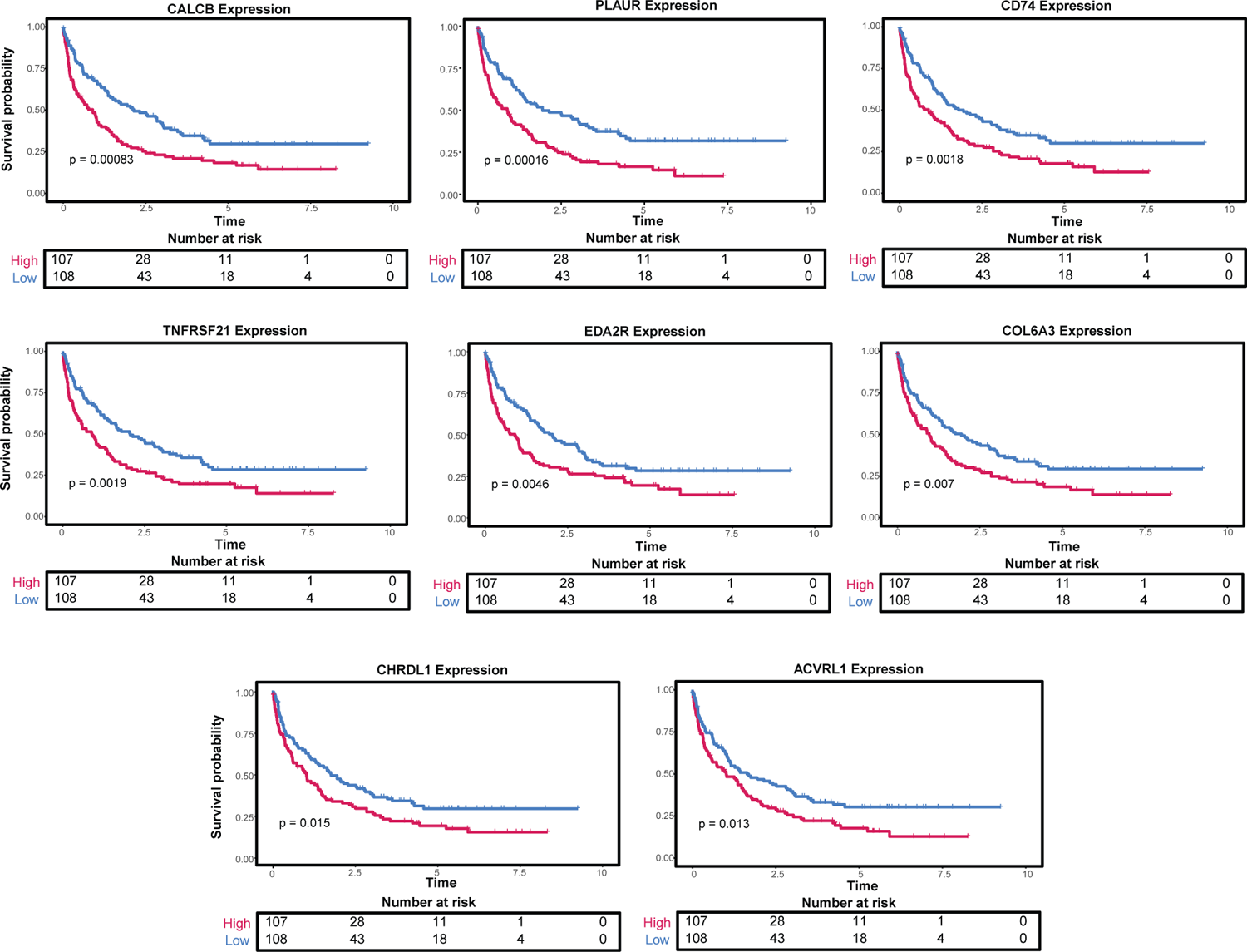
Overall survival using the 5-9 YBD cohort. Kaplan-Meier plots showing the association of each protein to OS.

## DISCUSSION

The identification of genetic variants associated with LC and their potential role in proteins levels in plasma can improve the understanding of the genetic predisposition for LC. Identifying circulating plasma proteins in individuals with a future diagnosis of LC can support the early detection of LC, improving the diagnosis and intervention of people with this disease. We used the data from the UKB to identify loci associated with LC and variants associated with the levels of proteins in plasma. To identify proteins that discriminate controls and patients with a future LC diagnosis up to 9 years after sample collection, we leveraged the proteome data found in the UKB-PPP. Finally, we used the identified proteins to classify samples taken after a LC diagnosis.

Performing a GWAS and a meta-analysis, we identified four loci that have not been previously reported to be associated with LC in previous studies and one locus that has been reported to be associated with LC in EA population but never in EUR (*9, 11*). We compared our results to the latest published LC meta-analysis (*31*) and found that the locus 10q24.3 previously reported to be associated with LUAD, was also associated with overall LC in our study. Interestingly, the gene *OBFC1/STN1* in this region of the genome has been reported to be involved in telomere regulation, an important mechanism in cancer (*36–40*).

We used the genome-wide significant variants from the UKB GWAS and the two meta-analyses to retrieve pQTLs associated with protein levels in plasma of individuals in the UKB-PPP. We found *cis*-pQTLs associated with higher levels of CTSH in all three studies. Pro-cathepsin H (CTHS) protein has an important role in the degradation of proteins in lysosomes, and high expression of this protein has been reported to be associated with favorable prognosis in thyroid carcinoma (*40*) and with tumor progression in prostate cancer (*41*). The role of this protein has not been investigated in LC. We also observed the positive association of rs6920364 with LC in the McKay *et al.* (*31*) meta-analysis, which is a lung *cis*-eQTL for *RNASET2*, and a plasma *cis*-pQTL associated with higher levels of RNASET2 in our study.

Ribonuclease T2 (RNASET2) is a ribonuclease that recognizes and degrades RNAs making it an important element in the innate immune system and has been associated as a tumor suppressor in ovarian cancer (*42*) and lymphomas (*43*) and a prognostic biomarker and therapeutic target in renal cell carcinoma (*44*). However, its role in LC needs to be further investigated.

In relation to the blood proteome, we established three cohorts consisting of cases and controls (0-4YBD, 5-9YBD, and 0-9YBD) using the data obtained from the UKB-PPP data. Using a logistic regression, we obtained 410, 340, and 807 DE analytes for each of the cohorts, respectively. When comparing our results to the biomarkers recently proposed by the Lung Cancer Cohort Consortium (LC3) (*27*), 30, 24 and 34 proteins out of 36 were also found to be DE in the 0-4YBD, 5-9YBD, and 0-9YBD cohorts, respectively. Of these proposed biomarkers, WFDC2, MMP12, GDF15, CEACAM5, CXCL17, and ALPP had the highest OR. Interestingly, the higher expression of WAP four-disulfide core domain 2 (WFDC2) in LUAD has been reported to play a role in the early detection of LC (*35*). The high expression of Macrophage metalloelastase 12 (MMP12) has been associated with chronic obstructive pulmonary disease (COPD), the transition from emphysema to LC and as a potential therapeutic target due to its role in tumor progression in the lung (*45*). Growth/differentiation factor 15 (GDF15) found in serum has been reported to be of significant value for the early diagnosis of LC and response to chemotherapy (*46*), and GDF15 plasma levels have been shown to have a predictive role in anti-PD-1/PD-L1 treatment in advanced LC (*47*). CEA-related cell adhesion molecule 5 (CEACAM5) has been reported to be highly expressed in LUAD and is being investigated as a potential therapeutic target (*48*), a predictive biomarker (*49*), and to play a role in the progression of NSCLC (*50*).

We identified the expression of eight proteins (EDA2R, PLAUR, CALCB, CD74, TNFRSF21, COL6A3, CHRDL1, and ACVRL1) to be associated with overall survival, where the high expression of these proteins is significantly associated with a shorter survival rate in samples taken up to 9 years before diagnosis. It is also worth mentioning that some of these proteins have been reported to potentially play roles in cancer. For example, the up regulation of Ectodysplasin A2 receptor (EDA2R) expression in tissue from lung carcinoma and melanoma mice has recently been reported to have a potential role in cancer cachexia (*51*), which has been associated with poor survival in NSCLC (*52*). High expression of HLA class II histocompatibility antigen gamma chain (CD74) has been associated as a prognostic biomarker and M1 macrophage infiltration marker in pan-tumor setting (*53*). CD74 has also been reported to be associated with the expression of PD-L1 in breast cancer and melanoma (*54, 55*). These findings suggest that CD74 may have a role in immunotherapy in multiple cancers, including NSCLC. Tumor necrosis factor receptor superfamily member 21 (TNFRSF21 or DR6) has been reported to promote necroptosis in osteosarcoma, accelerate tumor angiogenesis in B16 mice, and promote tumor aggressiveness in LC when it is overexpressed (*56–58*). DR6 has also been proposed as a diagnostic and predictive serum biomarker in adult sarcoma (*59*). Collagen alpha 3 (VI) chain (COL6A3) has been shown to be overexpressed in tumors compared to normal tissue in various cancers, including LC and proposed as a potential diagnostic and prognostic biomarker in cancer (*60*). In contrast to our findings, multiple studies have reported that a lower expression of Chordin-like protein 1 (CHRDL1) is associated with longer survival in NSCLC (*46, 61*). However, it is important to note that these studies primarily relied on RNA level data rather than protein level and were performed on different patient populations, which could potentially account for the conflicting results observed.

We identified the proteins that can effectively distinguish individuals with LC and control robustly. We demonstrated outstanding performance in classifying samples with a confirmed diagnosis, demonstrating relevance of the selected features to the disease. Specifically, when trained with the 0-4YBD, 5-9YBD, and 0-9YBD cohorts, the models achieved maximum AUCs of 0.81, 0.85, and 0.84 respectively. To assess the added value of proteins in LC risk assessment, we also trained ML models using only clinical variables such as age, sex, family history of LC, smoking status, and exposure to radon or asbestos. However, the highest AUC obtained with these models was 0.75. Previous studies have reported similar performance using the UKB-PPP data; however, these studies included all the incident participants into a single group (*62, 63*). In contrast, our 5-9YBD model achieved a notably high AUC, indicating its ability to accurately differentiate between individuals with lung cancer and controls while minimizing the inclusion of patients too close to the diagnosis time point.

To validate the proteins and findings obtained from the UKB models, we leveraged data from the LLP. In the three validation LLP cohorts, when the models were tested with samples gathered at the time of diagnosis, we observed AUCs only slightly lower than in the UKB-PPP models. Therefore, these outcomes reinforce the potential of the selected proteins to serve as reliable biomarkers for identifying individuals with lung cancer, even in diverse cohorts and datasets.

When we investigated the specific proteins utilized by the RF algorithm to achieve the observed performance, we observed that the RF algorithm relied on 209, 47, and 71 proteins in the 0-4YBD, 5-9YBD, and 0-9YBD cohorts, respectively. Among these proteins, we identified 22 that were found to be important across all cohorts. Previous studies have proposed 14 of these proteins (ALPP, CA6, CDCP1, CEACAM5, CXCL17, GDF15, IL15, LAMP3, PIGR, PLAUR, TNFRSF10B, TREM2, IFI30 and WFDC2) as potential biomarkers (*27, 62–64*). Interestingly, the remaining eight proteins have not previously been identified as potential biomarkers for early diagnosis of LC including ASGR2, CEACAM6, EDA2R, FASLG, IGDCC4, LRG1, NPC2, and TNR. Carcinoembryonic antigen-related cell adhesion molecule 6 (CEACAM6) has been reported to be overexpressed in multiple cancers including NSCLC, associated with a shorter survival and as a biomarker for leptomeningeal metastasis in LUAD (*65–67*). Tumor necrosis factor ligand superfamily member 6 (FASLG or CD95-L) has been associated with proliferation in multiple cancer types (*68*). NPC intracellular cholesterol transporter 2 (NPC2) has been shown to be secreted by pre-malignant tumor cells in mouse lung models, suggesting a role as a biomarker in early detection of LC (*69*). We also identified nine proteins that were only differentially expressed in incident cases, meaning they were uniquely associated with individuals who developed LC after blood collection (ALPP, ASGR2, CXCL17, FASLG, IGDCC4, IL15, LAMP3, TREM2, and TNR) and might play a role in the development of LC.

We have demonstrated that integrating pQTLs and genetic risk factors can increase the understanding of LC risk. Additionally, circulating proteins in plasma can be used to differentiate subjects who were diagnosed with LC up to 9 years after sample collection and controls. Although previous studies have suggested potential biomarkers in plasma, these were assessed up to 5 years from sample collection and including people with history of smoking (*27, 28, 32*).

Therefore, the strengths of our study include 1) the use of smokers and non-smokers in the cohort, which potentially captures biomarkers to support the screening of any individual at risk, 2) characterization of the proteome of individuals diagnosed with LC up to 9 years after sample collection, and 3) evaluation of the identified potential biomarkers using individuals with a confirmed LC diagnosis.

However, some of the limitations of our study include, 1) this study was done only using individuals of EUR ancestry and 2) histology and tumor stage were not available for all participants. Therefore, future cancer enhancement data and studies can expand our findings to other populations and evaluate whether the selected potential biomarkers are differentially expressed in different LC types and stages of the disease. Additionally, the identified proteins would need to be further evaluated experimentally to explore their potential role in LC development and progression and to confirm their ability to detect LC early.

In summary, our study revealed several significant findings. We identified four novel loci associated with LC in individuals of EUR ancestry, and one locus previously associated with LC in individuals of EA ancestry, but not in EUR populations. We investigated the relationship between the genome-wide significant variants and their associations with plasma protein levels. We showed that RNASET2 was associated with LC in two studies, with a lung *cis*-eQTL, and with a plasma *cis*-pQTL. Future studies will need to validate whether these pQTLs and LC associated variants share the same causal variant. Using the 2,937 analytes from the UKB-PPP study we successfully identified and externally validated proteins capable of classifying samples from patients with confirmed LC and controls, achieving AUC values greater than 0.80. Of the selected proteins, 233 exhibited a change in direction of effect post-diagnosis. Lastly, we identified a potential panel of 22 proteins to predict a future LC diagnosis.

In conclusion, our study provides valuable insights into the genetic predisposition to LC and the composition of the plasma proteome up to 9 years before an LC diagnosis in EUR populations, regardless of smoking status. These findings highlight potential genetic and proteomic factors associated with LC and contribute to our understanding of the disease.

## MATERIALS AND METHODS

### Genetics analysis UKB LC cohort

The UKB is a population-based, longitudinal, biomedical cohort consisting of over 500,000 individuals. It links demographic, clinical, environmental, lifestyle, and genetic information to better understand human health and diseases. Further details can be found at https://biobank.ndph.ox.ac.uk/showcase/.

To build a LC cohort, all the individuals of EUR ancestry in the UKB were selected (fields: 21000 and 22006, data version ukb674344). The 4,892 LC cases were selected using the UKB fields related to diagnosis or death (40001-40002, 41202-41205 and 41270-41271) and the International Classification of Diseases (ICD) 9 codes under 162 related to LC (162.2-162.5 and 162.8-162.9) and ICD-10 codes under C34 (C34.0-C34.3 and C34.8-C34.9). The 260,275 controls, were selected by filtering the rest of the EUR individuals (fields: 21000 and 22006) with the following criteria: 1) no record of any type of cancer (UKB fields: 41202-41205), 2) no individuals under ICD codes chapter II - malignant neoplasms (fields: 41270-41271), 3) cause of death not related to cancer (fields: 40001-40002), 4) no self-reported or prior history of cancer (field: 20001), and 4) no self-reported cancer medication (field: 20003). The following exclusion criteria was applied to both cohorts: 1) chr X aneuploidy (field: 22019), 2) mismatched biological and reported sex (fields: 31, 22001), 3) genetic outliers (field: 22027), and 4) individuals who have withdrawn their consent to be part of the UKB study by July 2023.

### GWAS with UKB LC cohort

The imputed single-nucleotide polymorphism (SNP) array data from the UKB LC cohort, containing individuals of EUR ancestry, was used to perform a GWAS using REGENIE v2.2.4 (*70*). The following variables were included as covariates: sex, ten genetic principal components (PC1-10), year of birth, smoking, and body mass index (BMI). Prior to plotting the results using the R package “CMplot” v.4.5.1 (*71*), SNPs were excluded from the summary statistics of the GWAS with the following criteria: 1) SNPs with a minor allele frequency less than 0.0001, 2) SNPs with an imputation score of less than 0.8, 3) SNPs with a Hardy-Weinberg Equilibrium *P* − *value* < 1*e*^-8^ in controls, and 4) SNPs with a missingness rate above 5%. After performing these filtering steps and selecting the significant variants (*P* − *value* < 5*e*^-8^), the results were compared to previously reported variants associated with LC in the GWAS catalog (*72*). Additionally, the variants were annotated using various tools including: the “variants to genes (V2G) score” available in Open Targets Genetics (*73, 74*), the combined annotation dependent depletion (*75–78*), the Ensemble Variant Effect Predictor (*79*), and the RegulomeDB (*33*).

### UKB and FinnGen Meta-analysis

The FinnGen study is a large-scale genomics initiative that has analyzed over 500,000 Finnish biobank samples and correlated genetic variation with health data to understand disease mechanisms and predispositions. The project is a collaboration between research organizations and biobanks within Finland and international industry partners (*30*).

Due to the similar phenotypic definition for cases and controls, the FinnGen summary statistics C3_BRONCHUS_LUNG_EXALLC (Release R11), “Malignant neoplasm of bronchus and lung (controls excluding all cancers),” were used to perform a meta-analysis with the UKB GWAS. A script in R v.4.3.2 was used to harmonize both summary statistics as following, 1) keep the variants available in both summary statistics, 2) check whether the alleles match for variants in common, if not, the variants were removed, 3) check the direction of effect for the matching alleles, if the direction did not match, we reversed the direction of effect in the FinnGen summary data. After harmonization, the files were used as input to METAL v. 2011-03-25 (*80*). Following METAL, the order of alleles and direction of effect were revised to be concordant with the LC UKB summary statistics, if they were flipped, the order and direction of effect were changed accordingly.

### UKB pQTL retrieval

The genome-wide significant variants (*P* − *value* < 5×10^-2^) from the UKB GWAS, UKB-FinnGen meta-analysis and the McKay *et al.* (*31*) were used to retrieve the significant pQTLs (*P* − *value* < 5×10^-2^) from the pQTLs reported in the UKB-PPP (*18*). Once the significant pQTLs were obtained, a one mega base window (1Mb) flanking the gene encoding the protein was utilized to define *cis* associations. pQTLs outside this window were considered as *trans*.

### Proteomics-based analysis

#### UKB-PPP case and control cohort selection

To further select participants who were diagnosed with LC during follow up, the proteomics data from the UKB-PPP was filtered using the participant ID numbers from the cohort used in the UKB GWAS analysis to determine cases and controls. The LC cases were divided into incident, those with an LC diagnosis after blood donation and prevalent, those with an LC diagnosis by the time of blood collection. The incident participants were divided into three categories, 0-4, 5-9 and 0-9 based on the years from blood donation until LC diagnosis. The normalized protein expression (NPX) of the 2,941 analytes representing 2,923 unique proteins from the UKB-PPP adjusted by technical confounders (batch, UKB center, array, blood season, and fasting time). This adjustment aimed to remove any potential biases introduced by these technical factors while retaining other nuisances that could be part of the disease.

Four proteins were excluded (PCOLCE, CTSS, GLIPR1, NPM1) due to having a missing rate > 20%. After filtering, missing values were imputed using the mean expression for each protein individually and 2,937 analytes representing 2,919 unique proteins were used for the analyses.

#### Differentially expressed plasma proteins in participants with a future LC diagnosis

The NPX technical and biological adjusted (batch, UKB center, array, blood season, fasting time, age, and sex) values of the proteins from the different categories were used as input to a logistic regression to obtain differentially expressed proteins between cases and controls. We determined significance at the threshold of *FDR* < 0.05 and an OR either greater than one for up regulated proteins or less than one for down regulated proteins.

#### Selection of proteins associated with LC in the different cohorts

The different cohorts were used to select proteins that differentiate cases from controls separately. Using each cohort, a random set of 70% was used to perform a 1000 iteration-bootstrapped logistic regression analysis using each of the NPX technical and biological adjusted (batch, UKB center, array, blood season, fasting time, age, and sex) values individually with smoking status as a covariate. Significant proteins were defined to be differentially expressed at an *FDR* < 0.05 in at least 100 of the bootstrapping iterations.

#### Predictive modeling

To compare the ability to classify prevalent cases with a confirmed LC diagnosis, six machine learning (ML) models were trained using a 5-fold cross-validation (CV) repeated 5 times. The models used were Logistic Regression (LogReg), ElasticNet (EN), XGBoost (XGB), Support Vector Machine (SVM) with a radial kernel, Random Forest (RF), and Multilayer Perceptron Neural Network (NN). The models were trained using the adjusted (batch, UKB center, array, blood season, and fasting time) NPX values of the selected proteins. Cases for each category and randomly selected controls were used in a 1:1 ratio as the training data for the models. Except for XGBoost (python XGBoost package v.1.7.3), all the models, were implemented using scikit-learn v.1.0.2 (*81*) in python. To evaluate the performance of the models, the AUC was used. The models were tested using prevalent cases and randomly selected controls (in a 1:1 ratio) from the controls that were not used during training. F1 scores were calculated function from the metrics module in scikit-learn. The RF models were used to retrieve the optimal features for each of the categories using the recursive feature elimination with cross-validation (RFECV) from scikit-learn (*81*).

#### Survival analysis using 5-9YBD cohort

To investigate the association between the levels of the 5-9YBD selected proteins and OS, a Cox proportional-hazards model was employed. The analysis was performed in R v.4.3.2 using the survival v.3.5-7 package (*82*). The LC baseline diagnosis date was determined using the UKB diagnosis fields (41270, 41280, 41271, 41281. For the time of event date, either the date of death (field: 4000), or the latest entry among the following categories was considered: date of lost follow up (field: 191), date of last personal contact with the UKB (field: 20143), or the last date of any diagnosis (fields: 41270, 41280, 41271, 41281). Proteins with a HZ greater than 1.3 and an FDR<0.05 were selected for further analysis. These proteins were used as input for a Kaplan-Meier estimator to examine the association between expression levels and OS. To conduct the Kaplan-Meier analysis, individuals were divided into two groups, low and high expression, based on the median expression level of each protein. The results of this analysis were then utilized to plot the Kaplan-Meier survival curve using the R package survminer v.0.4.9 (*83*).

#### Enrichment analysis of 5-9YBD proteins and pQTL-associated proteins

The 5-9YBD selected proteins and the pQTL-associated proteins were used as input to WebGestalt v.2019 (*84*) 47 separately to perform Gene Ontology (GO) (*85*) and pathway analyses. The pathway analysis included the Kyoto Encyclopedia of Genes and Genomes (KEGG) (*86–88*), Reactome (*89*), and Wikipathway (*90*) databases with the over-representation analysis (ORA) option. The GO analysis included biological process (BP), cellular components (CC), and molecular functions (MF). The 2,937 analytes from the UKB-PPP were used as reference and FDR < 0.05 was used to determine significance.

#### Independent validation

Using the longitudinal data from the LLP, the cohorts were established based on the diagnosis date of individuals, employing the same criteria used for the UKB-PPP analysis. The training cohort included individuals diagnosed with lung cancer (LC) within specific time intervals after the samples were taken. These intervals include less than 5 years, greater than 5 years but less than 9 years, or anytime between 0-9 years. The NPX values from the LLP were adjusted for storage time. This adjustment aimed to align the LLP NPX data with the normalization approach utilized in the UKB-PPP data used to train the ML models. New models were trained using the three LLP cohorts independently. Since the LLP contained longitudinal samples, individuals with multiple samples were only kept with the earliest sample collection. To test the performance of the models, the sample collection from the diagnosis date was used to build a new cohort of cases and healthy controls.

## Data Availability

This research has been conducted using the UK Biobank SNP Array Data and proteomic data under application numbers 52293 and 65851. The FinnGen R11 GWAS summary statistics on lung cancer was used for meta-analysis. Access to the UKB data can be requested through https://www.ukbiobank.ac.uk/enable-your-research/apply-for-access. Applications to access individual-level data in FinnGen can be made through the Finnish Biobanks’ “FinnBB” portal (https://finbb.fi/) and summary GWAS data can be accessed through the FinnGen website (https://www.finngen.fi/en/access_results).

## Acknowledgments

We would like to express our sincere gratitude to the participants and researchers of the UKB and FinnGen biobanks for their invaluable contributions in making the data available for our research. Furthermore, we would like to thank our colleagues, Drs. Iftekhar Kalsekar, Shuwei Li, and Katherina C. Chua for their valuable input and suggestions, which have significantly enriched our work.

## Author contributions

A.D.T., A.S, and M.A.J. contributed to the study’s design. L.H. preprocessed all the data. M.A.J. performed the data analyses and generated the Fig.s. All authors participated in the data interpretation. A.D.T. and A.S. jointly supervised this work. M.A.J. drafted the manuscript and all authors reviewed and edited the paper.

## Supplementary Materials

Fig. S1. Enrichment for GO, KEGG, Reactome, and Wikipathways of the 5-9YBD selected proteins

Fig. S2. Validation of UKB-PPP selected proteins using models trained with incident cases and testing with samples taken at diagnosis from the LLP study.

Table S1 to S4 Genetics Analysis

Table S5 to S6 Feature Selection

Table S7 Survival Analysis

Table S8 Potential Biomarkers

