## Supplementary Figures 1 and 2 for "Machine learning-based proteogenomic data modeling identifies circulating plasma biomarkers for early detection of lung cancer"

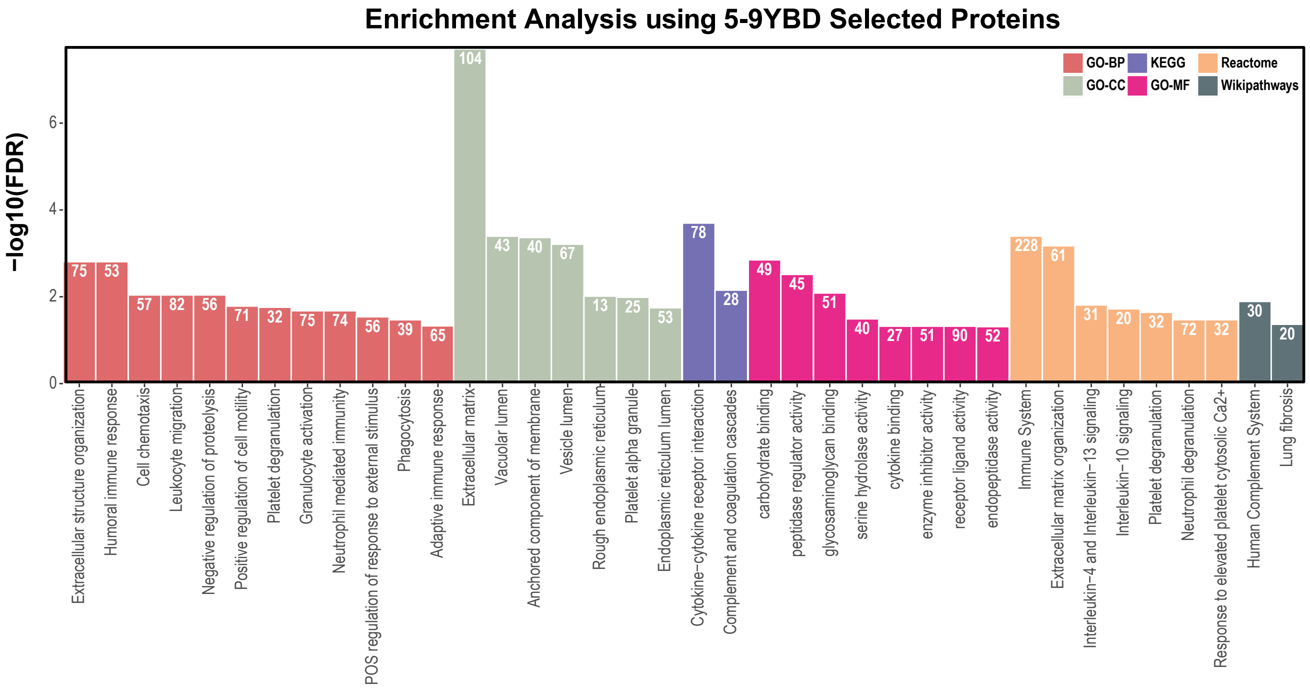


**Fig. S1. Enrichment for GO, KEGG, Reactome, and Wikipathways of the 5-9YBD selected proteins**. All pathways displayed achieved statistical significance with FDR threshold of less than 0.05. The number indicated above each bar represents the frequency of observed proteins associated with each pathway.

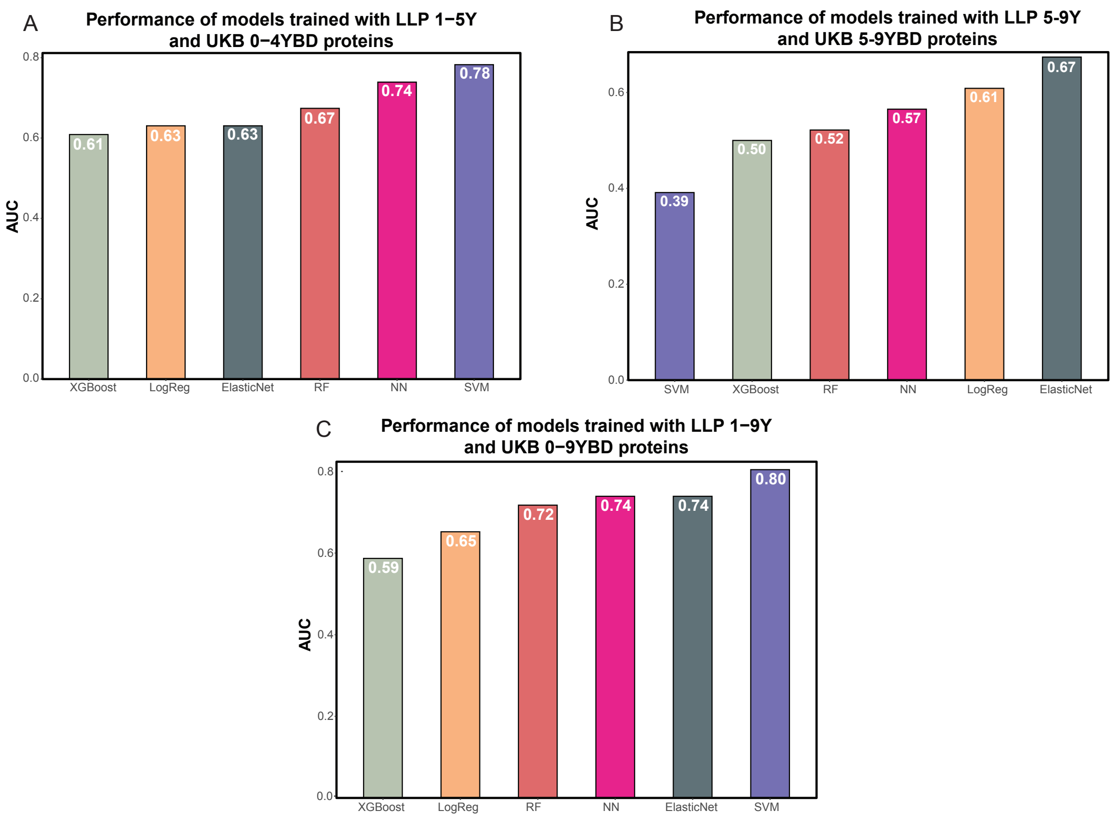


**Fig. S2. Validation of UKB-PPP selected proteins using models trained with incident cases and testing with samples taken at diagnosis from the LLP study.** A) AUC values obtained from the models trained on LLP 1-5Y cohort and the proteins selected from the 869 UKB 0-4YBD cohort. B) AUC values obtained when testing the models trained using the LLP 5-9Y cohort and the 782 proteins selected from the UKB 5-9YBD cohort. A) AUC values obtained when testing the models trained using the LLP 1-9Y cohort and the 491 proteins selected from the UKB 0-9YBD cohort.
